# A new tool to automate process map generation using Activity-Based Costing and Management data

**DOI:** 10.1101/2025.07.08.25328989

**Authors:** Joseph Corlis, Lori A. Bollinger, Kristin Bietsch

## Abstract

Activity-Based Costing and Management (ABC/M) examines the resources used, and their associated costs, to deliver health services through direct client observation. ABC/M has been supported in sub-Saharan Africa and south Asia by an array of funders to facilitate health program budgeting, monitoring, and sustainability planning. One output of ABC/M is the process map, which is a visual diagram that uses boxes, circles, arrows, and labels to depict how a health service is delivered. Process maps can illustrate the experience of a single client, including the time, location, order, and provider of each step in the delivery of a health service, or can show averages (e.g., the average time observed to deliver a health service) across clients at a specific health facility, across select facilities, or nationwide. Such process maps can be used by decision-makers at facilities as well as by national policymakers to identify inefficiencies, allocate resources, and/or explore adherence to protocols. Until now, generating process maps has been a key challenge, as ABC/M implementers had to create each separate process map manually, using shapes and text boxes in programs such as Microsoft PowerPoint, which was time-consuming and subject to error. The Analytics for Advancing the Financial Sustainability of the HIV/AIDS Response (AFS) project, with support from USAID and PEPFAR, developed code in R—an open-source, free programming language for statistical computing and data visualization—to automatically generate process maps. This code as well as instructions for users, legends for the graphics, and illustrative examples are all publicly available on GitHub. This article presents an illustrative set of process maps, which were developed for validity tests on the tool. Using this tool, researchers and policymakers can accurately construct each process map with data from ABC/M applications in minutes rather than days, as previously required.

## Introduction

Activity-Based Costing and Management (ABC/M) is a client-centered approach that can be used to identify resources used in health service delivery, inefficiencies, cost drivers, and possible areas for task-shifting, which can inform budget prioritization, sustainability planning, and facility monitoring. The methodology has been implemented in multiple countries with support from the United States Agency for International Development (USAID), the United States President’s Emergency Plan for AIDS Relief (PEPFAR), the United Nations Joint Programme on HIV/AIDS (UNAIDS), and other partners [1, 2]. ABC/M involves four main components: (1) tracking individual clients^1^ through each step of receiving a health service, from entry to exit, to determine the resources used during health services, described further below; (2) interviewing clients to capture their costs and experiences while accessing and using health services; (3) determining site-level overhead costs through desk review, key informant interviews, and focus group discussions; and (4) estimating above-site level costs for health services based on existing data. The first of these components is further defined as time-driven activity-based costing (TDABC). TDABC directly observes where clients go within a facility, the personnel they interact with, the duration of each step of the health service, which tests are ordered, and which resources are used (e.g., medicines, consumables, equipment) [3, 4].

ABC/M studies generally develop two types of process maps: normative and a combination of normative and empirical. Normative data represent the expected steps and characteristics of a health service visit based on standardized protocols by national agencies (e.g., Ministry of Health) as well as key informant interviews and focus group discussions with policymakers and clinical experts (e.g., health program director, hospital manager, technical working group member). Empirical data are gathered by study enumerators during the ABC/M study, and process maps that combine normative and empirical data use the normative steps and characteristics as a basis and layer on empirical data to demonstrate the actual steps and characteristics clients experience. The combination process maps can illustrate the experience of either a single individual or, more often, provide the average of all observed clients who received a specific health service, averaged across a single facility, a set of facilities, or nationally. Separate process maps must be created for each type of visit for a health service included in the study (e.g., first antenatal visit versus a follow-up visit), and they are commonly disaggregated further by client characteristics (e.g., newly diagnosed person living with HIV versus a virally suppressed client), health facility level (e.g., community clinic versus hospital) or geographic location (e.g., by province or region, rural versus urban) to account for attributes that may impact the resources available and flow of the service.

Process maps have a range of uses. Normative process maps can be used as visual guides for health service providers and trainees as a refresher on treatment protocols. Process maps that combine normative and empirical data can also be used for that purpose. Furthermore, decision-makers within facilities (e.g., clinical managers) could use the combination process maps as part of monitoring efforts to identify inefficiencies in time usage by providers or to monitor quality by identifying deviations from standard protocols. In addition, national level policymakers could use process maps that combine normative and empirical data to determine gaps in human resources within and across facilities, identify opportunities for task shifting in delivering specific health services, and inform resource allocation in budgets.

Until now, generating process maps has been a significant challenge for ABC/M studies, in terms of both the time required and ensuring data quality. Data analysts have often relied on multiple software programs to develop process maps, which is time and resource intensive, particularly for process maps that include empirical data. First, average time durations for each provider of each step of each service at each facility was calculated using statistical software. Next, a process map visual for each service for each facility, at either the individual or facility levels (or both), would be created in a program such as Microsoft PowerPoint with shapes that had to be individually arranged and color-coded. Then, text boxes would be overlaid on the arranged shapes to indicate the normative and empirical time durations and provider types for each step. Finally, each process map would be checked for data entry errors, which could arise at any point along the way. This time-consuming process often meant that only some of the possible process maps were ever created. In ABC/M reports, only a handful of key process maps are presented, usually showing facility averages of a small subset of services, but rarely showing individual level or regional averages, which could be useful for identifying quality issues or trends [5, 6, 7]. The new tool presented here addresses all these issues, including automating the calculations that underpin process map generation, quickly generating standardized initial process maps and subsequent revisions, and enabling stakeholders to craft new process maps based on emergent information needs without significant human resources requirements in training or time spent.

## Materials and Methods

### Ethics Statement

The development of this tool did not require ethical approval. Illustrative data were used during the creation and validation of the R code.

### Development of the tool and evaluation process

Developing this tool was identified at the beginning of the AFS project as critical for implementing ABC/M activities, based on lessons learned from previous ABC/M studies described above. During the tool’s development, we aimed to create a resource that could generate process maps automatically while also addressing four critical criteria: (1) minimize any required changes to the format of an existing ABC/M data set, (2) adhere to the standard format and symbols of process maps established by the Global Technical Committee on ABC/M, (3) be open source and freely available for all, and (4) require minimal training for users.

Previous ABC/M studies have collected data using different formats and media; some studies relied on paper forms that were later input into a statistical program for analysis while others collected data with specialized tablet-based software that could export the data in XLS or CSV format. We evaluated the data analysis templates used by previous ABC/M applications and found that, regardless of the medium for data collection, the data sets were generally structured in a similar way for analysis. We therefore structured our process map tool to accommodate XLS, CSV, and Google Sheets files that contain data organized such that each row indicates an individual client and columns indicate empirical data collected by enumerators during an ABC/M study, including the questionnaire number for each client, the facility where the client was observed, the region/district of the facility, the facility type, the health service visit observed, a brief description of the main task performed during each step of the health service visit, the start and stop times for each step of the health service visit, up to three providers participating in each step of the health service visit, and any laboratory tests that were ordered or received during each step of the health service visit. Both empirical and normative data should be entered using this format. By design, no personally identifying information can be input into the tool.

The Global Technical Committee for ABC/M previously established guidance that standardized the shapes, colors, symbols, and order for normative and combination normative-empirical process maps [3]. We followed this guidance (e.g., yellow circles in the bottom lefthand corner of each step signify the time for lab testing for each step) and created an automated legend that is attached to each process map. Because these shapes and symbols appear automatically, features related to empirical data may appear blank in combination process maps if no enumerators observed clients receiving a step (e.g., yellow circles in the bottom lefthand corner of a step only show numbers if a lab test was observed, grey circles in the bottom righthand corner of a step only show numbers if a client received the step).

Beyond the established guidance, our tool expands the process maps to include more information than previous process maps have shown. For example, in combination normative-empirical process maps, we added a bar graph below each step to indicate the distribution of providers who *actually* participated (as percentages) rather than color-code each step with the provider who *should* participate based on norms as previous process maps have. Additionally, our tool produces process maps that indicate the average wait time between steps.

We excluded one design element from process maps, which are included in the established guidance— separate shapes representing decision nodes—for two reasons. First, prior applications often included decision nodes for only some of the decisions that providers must make over the course of a client’s health service visit (e.g., whether to conduct an HIV viral load test during a visit for antiretroviral therapy) while excluding decision nodes for other decisions that may arise, particularly in integrated health services (e.g., whether to offer multiple services in one visit). For consistency, we would need to include decision nodes for all possible decisions, and that would make the process maps very detailed and complex, reducing their utility. Alternatively, the decision nodes would need to be individually tailored for each process map and would reduce the flexibility and efficiency that was the original intention of creating this tool. To compensate for this, while we excluded decision nodes, we added the percentage of clients receiving each step (indicated above the arrow between steps) to combination normative-empirical process maps to ensure no information is lost.

To reduce barriers to use related to price or lack of skills, we developed the tool using R, a free software program with an online user support community should general technical questions arise. The legends, which we also created in R, can be updated using R software or by editing the legend graphics in programs that permit editing PNG files (e.g., Canva, Microsoft Paint, Microsoft PowerPoint). Our tool, which includes the R code, pre-made legend graphics, CSV files containing illustrative data, illustrative final process map examples, and a full set of complete instructions, are all available at https://github.com/AvenirHealth-org/ProcessMaps.

We conducted several rounds of validity tests of the R code, iterating the code and corresponding materials internally and then aligning the tool with feedback from USAID and the Global Technical Committee on ABC/M.

### Example Application and Analysis

To ensure that the tool produces accurate process maps, we constructed a practical application of the process map code based on ABC/M applications conducted under projects funded by USAID, the Gates Foundation, and other donors. Since data from ABC/M applications are often proprietary, we generated an illustrative data set in Microsoft Excel for 50 hypothetical individuals receiving antiretroviral therapy (ART) services for a new diagnosis of HIV. These 50 people living with HIV (PLHIV) either received treatment at a public district hospital or a private hospital. Figure 1 shows the structure of the data set and a portion of the illustrative data. We entered both normative data related to treatment standards for newly diagnosed PLHIV on ART as well as illustrative data for the 50 individuals, then we uploaded the data set into the R code as an XLS file.

**Figure 1:**
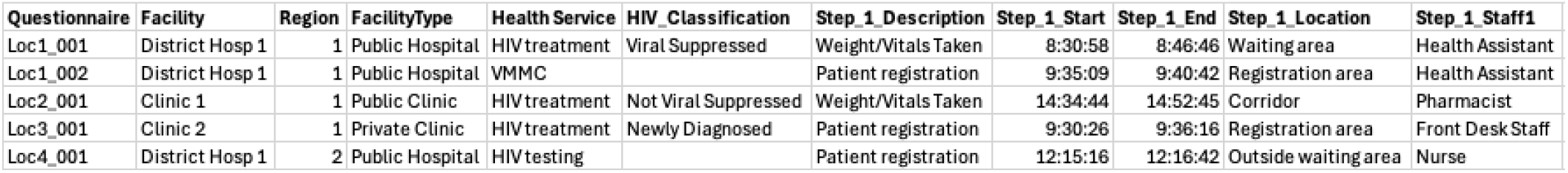
Data structure template for input into the R code for process map generation with illustrative data.

We used the R code to generate several process maps based on the illustrative data set. Figure 2 shows a comparison of the client averages at a district public hospital as well as the normative values that such a hospital might expect for step duration, provider type, and step location during a newly diagnosed PLHIV’s ART service visit. Similar figures can be generated for each facility, thus allowing for comparisons both across facilities and between facility averages and norms. For example, Figure 2 shows that not all clients at the public district hospital received every step of ART expected in the normative client flow for a hospital (e.g., less than 30% of clients received nutritional assessments or had blood samples taken), and clients had longer wait times than the normative process. Nurses and health assistants had the most contact with clients at the public district hospital, conducting steps expected for doctors in the normative process map.

**Figure 2:**
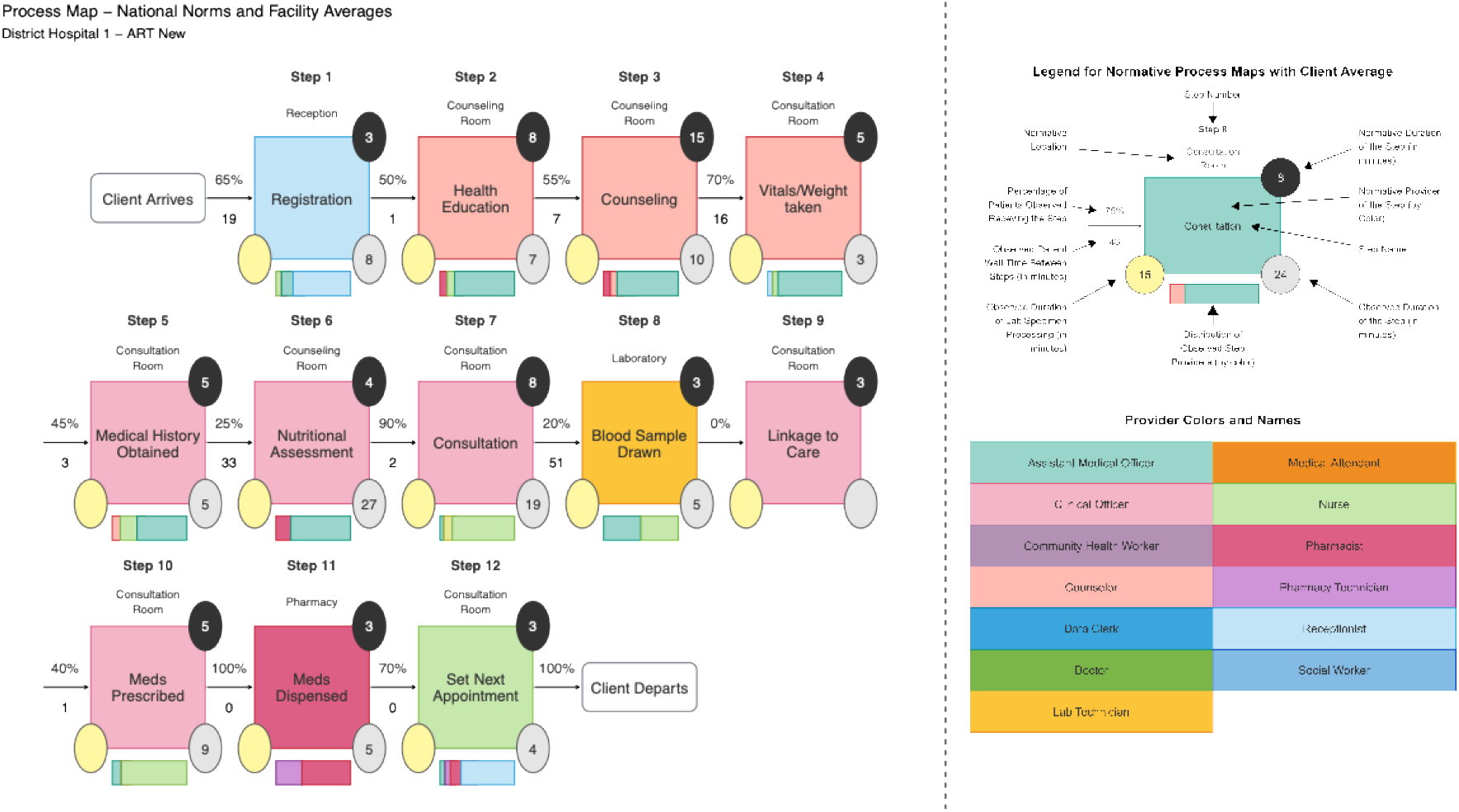
Example process map generated in R that compares national norms of HIV treatment services for newly diagnosed clients with illustrative client averages for two hospitals (left), along with the legend for the process map (right).

Figure 3 illustrates another type of process map that the R code can generate: an average of the illustrative empirical data for the public district hospital compared to the normative processes for a hospital (as in Figure 1) in the top panel with individual client-level process maps beneath. This figure shows that the average time in Normative Step 7 (consultation), which far exceeds the national norms, is driven largely by the experience of clients 14, 17, and 20 whose observed time in consultation were more than triple the norm. In addition, in this step these clients were also seen by student nurses instead of nurses or doctors like the other clients. Personnel costs for student nurses are lower than for nurses or doctors, which means that the total expense for clients 14, 17, and 20 may not differ significantly from the average expenditure for the total health service visit at the district public hospital. Nevertheless, Figure 3 reveals how much longer these clients spent receiving care than the average client at the hospital and how divergent their experience was from national norms.

**Figure 3:**
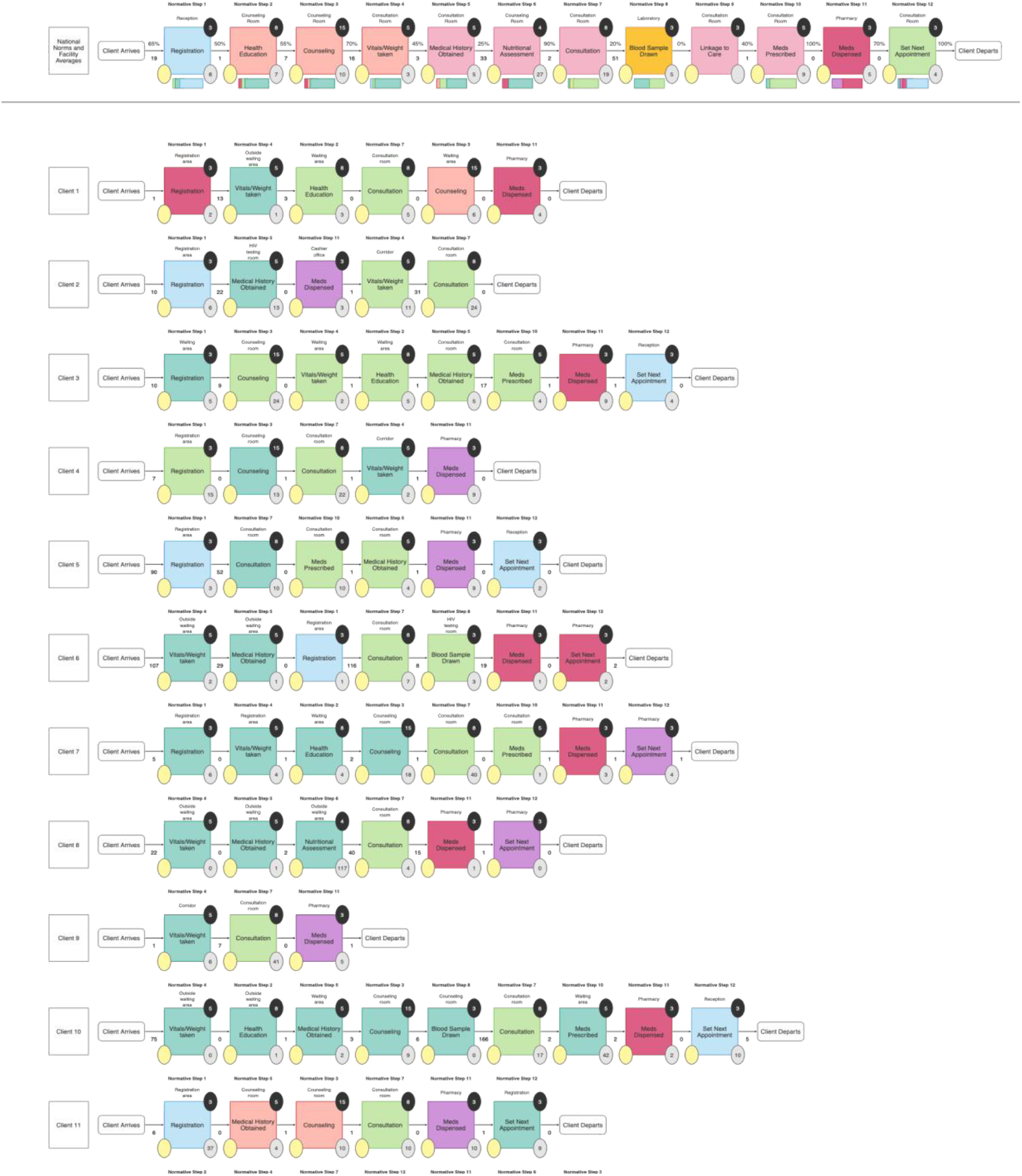
Example process map generated in R for HIV treatment services for newly diagnosed clients that includes a comparison of national norms and facility-level averages for a representative public district hospital (top panel) and illustrative client-level observations used in the averages (bottom panel).

Note that, in both Figure 2 and Figure 3, blank yellow and grey circles attached to some steps indicate that no labs or provider interactions were observed for that step, even if the steps are expected in the normative process for the health service.

### Implementation methods

Using the tool requires access to a PDF reader (to view the instructions), RStudio (to run the code)^2^, and a data entry software such as Microsoft Excel (to enter, format, and export data to be uploaded to RStudio). If the process map legends need to be modified, a graphic design software such as Microsoft Paint, Microsoft PowerPoint, or Canva may be used instead of RStudio.

This tool does not affect the use cases for process maps. Rather, this tool was conceived to make creating the visuals easier and faster. As with process maps developed for previous ABC/M studies, the outputs of this tool can be disseminated based on the decisions of the country-led ABC/M steering committee [4].

## Discussion

Given the well-documented global gaps in resource allocation and cost data, ABC/M has emerged as an innovative methodology for determining resource use and cost information [8, 9]. Other resources have been developed to guide implementers and policymakers in generating and using ABC/M data to improve resource allocation decision-making and efficiency for health programs, including sample research protocols and data collection instruments [4, 10]. Our new tool is the first resource to automate the generation of process maps, addressing a key gap identified in previous ABC/M applications. This tool, in conjunction with other ABC/M guidance documents, can provide critical information on resources used at the client-, facility-, regional, or national level. To date, the absence of such an automated tool has limited the utility of process maps to guide the decision-making of public health implementers and officials.

Our tool currently faces two main limitations. First, the tool includes generic names for health providers and spaces within health facilities, which may not accurately match the names in every context. Updating these names in the process map legends is possible, though novice R coders may find this difficult. Second, the tool is currently only available in English. While the process map outputs could be translated into other languages by updating the R code and legends, most users would likely find this tedious. Future updates to the tool aim to include language translation capabilities.

Further refinements to this tool could be made based on lessons learned while implementing future ABC/M studies. In addition, feedback on the usefulness of the tool’s outputs by country-based and global technical working groups engaged in decision-making related to health program costs, financing, and resource usage is most welcome. Future iterations of this tool may also reflect ongoing efforts to integrate vertical programs into existing health service delivery systems [11, 12, 13]. In summary, by drastically reducing the time necessary to generate accurate and timely process maps, we hope this tool will serve as a public good that can accelerate discussions of health program efficiency and sustainability across a wide variety of applications.

## Data Availability

All data produced in the present study are available upon reasonable request to the authors.

## Acknowledgements

The authors wish to acknowledge the feedback that USAID and the Global Technical Committee on ABC/M provided during the development of this process map tool.

## Footnotes

1 The term *client* is used to indicate anyone who receives a health service or product; in many contexts, such individuals may also be referred to as a *patient, beneficiary, service user, end user*, or other similar term.

2 Available at https://posit.co/download/rstudio-desktop/.

